# Age-Stratified Patterns of Diagnostic Delay in Pertussis

**DOI:** 10.64898/2026.09.25.26364016

**Authors:** Emily A. Serman, Iromi Jayawardena, Carolina Liskey, Robin Lacroix, Alain H. Litwin, Jihad S. Obeid, Scott R. Curry, Lior Rennert

**Affiliations:** Department of Public Health Sciences, Clemson University, Clemson, SC, USA; Center for Public Health Modeling and Response, Clemson University, Clemson, SC, USA; Medical University of South Carolina, Charleston, SC, USA; University of South Carolina School of Medicine-Greenville, Greenville, SC, USA; Prisma Health-Upstate Children’s Hospital, Greenville, SC, USA; Prisma Health-Upstate, Greenville, SC, USA; Division of Infectious Diseases, Medical University of South Carolina, Charleston, SC, USA

**Keywords:** Consumer engagement, Precision medicine, Delphi study

## Abstract

**Background:** Pertussis has resurged in the post-COVID-19 era, yet diagnostic delay remains a barrier to timely treatment and transmission interruption, particularly among adults.

**Methods:** Retrospective observational cohort study of 787 patients with confirmed pertussis from January 2016 through March 2026 across two South Carolina health systems. Electronic health record data from the 60 days preceding confirmation were used to characterize diagnostic timing (same-day vs. delayed), delay duration, and respiratory pathogen testing patterns including pertussis test modality.

**Results:** Adults accounted for 48.4% of cases (median age 16 years [IQR 4–52]; 61.6% female). Same-day diagnosis was highest among children aged 1–4 years (75.4%) and lowest among adults aged 50 and older (approximately 50%). Among the 305 patients (38.8%) with delayed diagnosis, median delay ranged from 7–12 days in children to 22.5–26.5 days in adults aged 50 and older. Delay did not vary significantly by COVID-19 period, race, ethnicity, or insurance type. Testing breadth was greater in inpatient and emergency settings than outpatient. Pertussis testing was predominantly PCR-based in children but serologic in adults, potentially reflecting later clinical presentation relative to illness onset; however, the near-absence of PCR testing in older adults may contribute to missed diagnoses during the early illness window when PCR sensitivity is highest.

**Conclusions:** Older adults experienced markedly delayed pertussis diagnoses compared with children. Earlier consideration of pertussis in adults with prolonged cough, including PCR testing early in illness before serologic conversion, may reduce diagnostic delay and facilitate timelier treatment and contact tracing.

**Summary:** Among 787 confirmed pertussis patients across two health systems, adults aged 50 and older experienced diagnostic delays two to three times longer than children, likely driven by atypical symptom presentation and late-stage testing patterns that fall outside optimal diagnostic windows.

## Introduction

Bordetella pertussis, the causative agent of pertussis (whooping cough), produces a characteristic paroxysmal cough that in infants and young children is often accompanied by an inspiratory whoop [1]. Pertussis follows a defined three-stage clinical course − catarrhal, paroxysmal, and convalescent − each lasting one to two weeks, with the catarrhal phase most closely resembling other common upper respiratory illnesses and therefore most vulnerable to diagnostic error [1, 2]. The disease has long been recognized as a leading cause of vaccine-preventable mortality in children under one year of age [1]; however, clinical presentation varies substantially across the lifespan. While young children may present with classic paroxysmal cough, cyanosis, and post-tussive emesis, infants, older children and adults frequently exhibit attenuated or atypical symptoms that often do not prompt immediate clinical suspicion for pertussis [2, 3, 4, 5]. As one of the most contagious respiratory pathogens, with attack rates of approximately 90% in household contacts and 50–80% in classroom settings [6], delayed or missed pertussis diagnosis carries significant implications for transmission and patient outcomes.

Timely diagnosis is critical on two fronts: antibiotic treatment is most effective when initiated during the catarrhal phase within the first one to two weeks of symptom onset [2], and rapid identification enables isolation and contact prophylaxis to interrupt transmission [7]. This urgency has intensified in the post-COVID-19 era. After a sharp pandemic-era decline, attributed to non-pharmaceutical interventions and reduced healthcare utilization, pertussis has resurged globally [8, 9, 10, 11, 12, 13], with U.S. cases increasing five-fold in 2024 compared to 2023 and prompting state health alerts [6, 14, 15]. This resurgence is compounded by declining childhood vaccination rates and geographic clustering of vaccine exemptions [6, 16, 17], and further complicated by the overlapping respiratory symptom profiles of COVID-19 and pertussis, which may distort clinical recognition and extend delays beyond pre-pandemic estimates.

Prior research has documented diagnostic delays across age groups. Most notably, Evans et al. (2023), in the largest study of pertussis diagnostic delay to date using insurance claims data from over 20,000 patients, found that delays occurred in 29.7–37.6% of cases, with duration increasing considerably with age − from an average of 5.6 days in patients under two years to 13.8 days in adults [18]. However, existing literature is limited in several respects. Many prior studies focus narrowly on pediatric populations, and even those spanning the full age spectrum do not separate infants from toddlers aged one to four despite well-documented differences in clinical presentation between these groups and the potential implications for diagnostic recognition [2]. Furthermore, diagnostic testing patterns, including testing breadth and laboratory modality, have not, to our knowledge, been characterized in prior work, despite their potential to illuminate why delays occur. Finally, no study to our knowledge has characterized how diagnostic delay has changed across the pre- and post-COVID-19 periods, when both case burden and clinical context have shifted considerably.

This study examines patterns of diagnostic delay in pertussis across two health systems, characterizing prior healthcare encounters, diagnostic misattribution, testing patterns, and time-to-diagnosis by age group, sex, race and ethnicity, insurance type, and epidemiological period.

## Methods

### a. Data

This study used Electronic Health Record (EHR) data from two of South Carolina’s largest health systems: a large community-based health system covering approximately 1.4 million patients annually across a 26-county service area, and a statewide academic medical center serving 46 counties with approximately 3.21 million patient encounters annually. These health systems are hereafter referred to as Health System A and Health System B. De-identified data were extracted from the EHR, including inpatient, emergency department, and outpatient encounter records, diagnosis codes, admission and discharge dates, and laboratory results. This study was approved by the Institutional Review Board at Clemson University (IRB2024-0015).

### b. Study Design

This was a retrospective observational study conducted across two health systems in South Carolina spanning the period January 2016 through March 2026. Individuals with pertussis were defined as those with a pertussis-specific International Classification of Diseases, Tenth Revision (ICD-10) diagnosis code (eTable 1) or a positive laboratory test for Bordetella pertussis, including PCR or serologic testing (IgG, IgA, or IgM). For each patient, we examined respiratory-related healthcare encounters within a 60-day window preceding the index pertussis date, selected to capture the typical duration of symptomatic pertussis from catarrhal presentation through diagnosis.

**Table 1:** Demographic and clinical characteristics of the study cohort by health system.

| **Characteristic (n (%))** | **Overall**<br>**N = 787** | **Health System**<br>**A**<br>**N = 305** | **Health System**<br>**B**<br>**N = 482** | **p-**<br>**value<sup>a</sup>** |
| --- | --- | --- | --- | --- |
| **Age (years), median (IQR)** | 16.0 (4.0, 52.0) | 18.0 (5.0, 54.0) | 15.0 (3.0, 51.0) | 0.2 |
| **Age group** |  |  |  | 0.5 |
| <1 (infant) | 99 (12.6%) | 33 (10.8%) | 66 (13.7%) |  |
| 1-4 | 126 (16.0%) | 43 (14.1%) | 83 (17.2%) |  |
| 5-17 | 181 (23.0%) | 75 (24.6%) | 106 (22.0%) |  |
| 18-49 | 170 (21.6%) | 66 (21.6%) | 104 (21.6%) |  |
| 50-64 | 95 (12.1%) | 43 (14.1%) | 52 (10.8%) |  |
| 65+ | 116 (14.7%) | 45 (14.8%) | 71 (14.7%) |  |
| **Sex** |  |  |  | 0.8 |
| Female | 485 (61.6%) | 186 (61.0%) | 299 (62.0%) |  |
| Male | 302 (38.4%) | 119 (39.0%) | 183 (38.0%) |  |
| **Race** |  |  |  | 0.090 |
| White | 564 (71.7%) | 211 (69.2%) | 353 (73.2%) |  |
| Black | 110 (14.0%) | 53 (17.4%) | 57 (11.8%) |  |
| Other | 113 (14.4%) | 41 (13.4%) | 72 (14.9%) |  |
| **Ethnicity** |  |  |  | >0.9 |
| Hispanic | 72 (9.1%) | 27 (8.9%) | 45 (9.3%) |  |
| Non-Hispanic | 696 (88.4%) | 271 (88.9%) | 425 (88.2%) |  |
| Unknown/Refused | 19 (2.4%) | 7 (2.3%) | 12 (2.5%) |  |

| Characteristic (n (%)) | Overall<br>N = 787 | Health System<br>A<br>N = 305 | Health System<br>B<br>N = 482 | p-value<sup>a</sup> |
| --- | --- | --- | --- | --- |
| **Insurance type** |  |  |  | <0.001 |
| Self-Pay/Uninsured | 205 (26.0%) | 14 (4.6%) | 191 (39.6%) |  |
| Medicaid | 163 (20.7%) | 61 (20.0%) | 102 (21.2%) |  |
| Medicare | 113 (14.4%) | 45 (14.8%) | 68 (14.1%) |  |
| Commercial/Private | 271 (34.4%) | 154 (50.5%) | 117 (24.3%) |  |
| Other | 4 (0.5%) | 3 (1.0%) | 1 (0.2%) |  |
| Missing | 31 (3.9%) | 28 (9.2%) | 3 (0.6%) |  |
| **COVID-19 period** |  |  |  | 0.9 |
| Pre-COVID-19 (2016-2019) | 246 (31.3%) | 96 (31.5%) | 150 (31.1%) |  |
| COVID-19 period (2020-2023) | 131 (16.6%) | 48 (15.7%) | 83 (17.2%) |  |
| Post-COVID-19 (2024-2026) | 410 (52.1%) | 161 (52.8%) | 249 (51.7%) |  |
| **Diagnostic timing** |  |  |  | 0.013 |
| Delayed | 305 (38.8%) | 135 (44.3%) | 170 (35.3%) |  |
| Same-day | 482 (61.2%) | 170 (55.7%) | 312 (64.7%) |  |
| **Prior encounters before diagnosis** |  |  |  | 0.041 |
| 0 (same-day) | 482 (61.2%) | 170 (55.7%) | 312 (64.7%) |  |
| 1 | 159 (20.2%) | 66 (21.6%) | 93 (19.3%) |  |
| 2 | 74 (9.4%) | 32 (10.5%) | 42 (8.7%) |  |
| 3+ | 72 (9.1%) | 37 (12.1%) | 35 (7.3%) |  |

<sup>a</sup>Wilcoxon rank sum test; Fisher's Exact Test for Count Data with simulated p-value (based on 10000 replicates); Fisher's exact test

### c. Classification of Diagnostic Timing

Patients were classified as having a same-day diagnosis if their index pertussis date coincided with their first respiratory-related healthcare encounter within the 60-day lookback window, where respiratory-related encounters were defined using the ICD-10 code categories described in eTable 2. Patients were classified as having a delayed diagnosis if at least one respiratory-related healthcare encounter was identified prior to the index pertussis date within the 60-day window. For patients with delayed diagnoses, the duration of delay was defined as the number of days elapsed between the first qualifying prior encounter and the index pertussis date.

**Table 2:** Diagnostic delay rate and duration by age group and health system, based on a 60-day lookback window.

|  | **Overall** |  | **Health System A** |  | **Health System B** |  |  |
| --- | --- | --- | --- | --- | --- | --- | --- |
| **Age Group** | **Delayed, n/N (%)** | **Median Days<sup>a</sup>** | **Delayed, n/N (%)** | **Median Days<sup>a</sup>** | **Delayed, n/N (%)** | **Median Days<sup>a</sup>** | **p-value<sup>b</sup>** |
| <1 years (infant) | 36/99 (36.4%) | 11 | 12/33 (36.4%) | 11 | 24/66 (36.4%) | 10.5 | 0.260 |
| 1-4 years | 31/126 (24.6%) | 7 | 12/43 (27.9%) | 5 | 19/83 (22.9%) | 9 | 0.744 |
| 5-17 years | 64/181 (35.4%) | 12 | 27/75 (36%) | 14 | 37/106 (34.9%) | 11 | 0.177 |
| 18-49 years | 68/170 (40%) | 20.5 | 35/66 (53%) | 19 | 33/104 (31.7%) | 21 | 0.927 |
| 50-64 years | 48/95 (50.5%) | 26.5 | 27/43 (62.8%) | 27 | 21/52 (40.4%) | 26 | 0.942 |
| 65+ years | 58/116 (50%) | 22.5 | 22/45 (48.9%) | 21.5 | 36/71 (50.7%) | 23.5 | 0.860 |
| **Overall<sup>c</sup>** | **305/787 (38.8%)** | **18** | **135/305 (44.3%)** | **19** | **170/482 (35.3%)** | **17** | N/A |

<sup>a</sup>Median days to diagnosis among delayed patients only.
<sup>b</sup>Wilcoxon rank sum test comparing delay duration between health systems among delayed patients only.
<sup>c</sup> $\chi^2(5) = 23.6$ , $p < 0.001$ for difference in delay rates across age groups.

### d. Covariates

Key demographic and clinical covariates were obtained from patient records within the EHR. Age was categorized into six groups: less than 1 year (infants), 1–4 years, 5–17 years, 18–49 years, 50–64 years, and 65 years and older. Sex was recorded as male or female. Race was categorized as White, Black, or Other. Ethnicity was categorized as Hispanic or Non-Hispanic. Insurance type was defined using payer information recorded at the index encounter, or nearest encounter if needed, and categorized as Commercial/Private, Medicaid, Medicare, or Self-Pay/Uninsured. Cases were additionally stratified by epidemiological period: pre-COVID-19 (2016–2019), COVID-19 period (2020–2023), and post-COVID-19 resurgence (2024–present). Laboratory testing data were extracted from EHR records within the lookback window. Respiratory pathogen testing breadth was quantified as the number of distinct pathogens tested per encounter. Pertussis-specific tests were classified by modality as PCR, Serology (IgG, IgA, or IgM), or Other (unspecified), based on laboratory order names. Testing patterns were examined by care setting, age group, and diagnostic timing.

### e. Statistical Analysis

Differences in the proportion of patients with same-day versus delayed diagnosis across demographic and temporal subgroups were assessed using Pearson chi-square tests. Where the omnibus chi-square was significant, pairwise proportion tests were conducted with Bonferroni correction for multiple comparisons. Among patients with delayed diagnoses, differences in delay duration were assessed using Kruskal-Wallis rank sum tests given the right-skewed distribution of delay times, with Dunn’s post-hoc test applied for pairwise comparisons where the omnibus test was significant. As a complementary multivariable approach, we fit a discrete-time complementary log-log hazard model to estimate the adjusted hazard of same-day diagnosis, with each patient’s follow-up divided into person-day observations up to a 60-day horizon (with the diagnosis day counted as day 0). Baseline hazard was modeled using six time bands (day 0; days 1-2; 3-7; 8-14; 15-30; and 31-59) to allow flexible, non-linear variation in diagnostic hazard over time. The model included age group, sex, race, ethnicity, insurance type, health system, and epidemiological period as covariates, with hazard ratios (HRs) and 95% confidence intervals obtained by exponentiating model coefficients. All analyses were conducted in R (version 4.5.1). Statistical significance was defined as p < 0.05.

## Results

### a. Overview

Of 787 patients with confirmed pertussis from 2016 through March 2026, 305 were from Health System A and 482 from Health System B (Table 1). The two systems were comparable in age, sex, race, ethnicity, and COVID-19 period (all p>0.05) but differed significantly in insurance mix (p<0.001), with Health System A serving more commercially insured patients (50.5% vs. 24.3%) and Health System B serving more self-pay/uninsured patients (39.6% vs. 4.6%). The proportion of delayed diagnoses was higher at Health System A than Health System B (44.3% vs. 35.3%; p=0.013), though delay duration was comparable across sites (Wilcoxon p=0.176). The primary age-related gradient in diagnostic delay was consistent across both systems; combined analysis is therefore presented as primary, with system-stratified results in Table 2.

### b. Same-Day vs. Delayed Diagnosis

The proportion of same-day versus delayed diagnoses differed significantly across age groups (χ^2^(5) = 23.6, p<0.001). The 1–4 year age group had the highest same-day diagnosis rate at 75.4%, while adults aged 50-64 and 65+ had the lowest with both around 50%. Pairwise comparisons confirmed significant differences between the 1–4 group and both the 50–64 group (p = 0.002) and the 65+ group (p = 0.001) after Bonferroni correction. Table 2 presents the full diagnostic timing summary by age group and health system. Delayed-diagnosis rates did not differ significantly across the epidemiological periods overall (chi-square p=0.242), though rates among adults aged 50 and older remained persistently elevated across all three periods (eTable 3).

### c. Duration of Diagnostic Delay

Among the 305 patients with delayed diagnoses, the duration of delay differed significantly across age groups (Kruskal-Wallis H=14.7, df=5, p=0.012). Median delay was shortest among children aged 1–4 years and 5–17 years (7 and 12 days, respectively) and longest among adults aged 50–64 years and 65+ (26.5 and 22.5 days), with adults aged 18–49 showing an intermediate delay of 20.5 days (Table 2). Infants showed a median delay of approximately 11 days. No individual pairwise comparison reached significance after Bonferroni correction, consistent with a broad age gradient rather than discrete group differences. Diagnostic timing differed by sex, with female patients having a higher proportion of delayed diagnoses than male patients (41.6% vs. 34.1%; chi-square p=0.042); however, the duration of delay did not differ significantly by sex (median 19 days females vs. 17 days males; Wilcoxon p=0.526). Diagnostic timing did not differ significantly by race, ethnicity, or insurance type (all chi-square p>0.05); delay duration was similarly non-significant across race (Kruskal-Wallis p=0.412) and ethnicity (Wilcoxon p=0.125); full descriptive statistics are provided in eTable 4. The Kruskal-Wallis test for delay duration by insurance type was significant (p=0.016), and Medicare patients showed a longer median delay than Medicaid patients (27.5 vs. 8.5 days; Dunn post-hoc p=0.007). Prior encounter diagnoses were predominantly respiratory across all age groups and consistent with established age-related presentation differences, with upper respiratory infection and cough predominating in young children and shortness of breath and pneumonia leading among adults 50 and older, likely contributing to the longer delays observed in this group (eTables 5 and 6). Age was a significant predictor of length of delay in the adjusted discrete-time hazard model (eTable 7); Sex, race, ethnicity, insurance type, and epidemiological period were not statistically significant predictors.

### e. Respiratory Pathogen Testing Patterns

Laboratory testing data were available for 589 of 787 cohort patients (74.8%); the remaining 198 patients (25.2%) had no laboratory testing documented, of whom 184 (92.9%) received a same-day pertussis diagnosis. Pathogens tested per encounter differed significantly by care setting (Kruskal-Wallis p<0.001), with all pairwise comparisons significant after Bonferroni correction. Inpatient encounters involved the broadest testing (mean 7.2 pathogens per encounter), followed by emergency department encounters (mean 5.6), and outpatient encounters (mean 4.0); consistent across both health systems (eTable 8).

Testing breadth also differed significantly by age group (Kruskal-Wallis p<0.001). Infants and children aged 1–4 were tested most broadly (mean 6.2 and 5.9, respectively), while adults aged 18–49 and 65+ were tested least (mean 3.4 and 3.5, respectively), a pattern that was most pronounced in the outpatient setting. Full age- and setting-stratified data are presented in eTables 9 and 10.

Testing breadth did not differ significantly between same-day and delayed patients overall or within either health system individually (all p>0.05), suggesting that testing breadth alone does not determine diagnostic timing (eTable 11). However, diagnostic timing differed significantly by care setting at the index encounter (χ^2^(2)=34.7, p<0.001): delayed diagnosis was most common when the index encounter occurred in the outpatient setting (41.9%) and least common in the emergency department (15.6%) (eTables 12 and 13).

### f. Pertussis-specific Testing Patterns

Pertussis test type varied by age group (eTable 14). PCR predominated in infants and children aged 1–4 at both prior and index encounters (83.8% and 72.4% of tests at index, respectively). In adults, serologic testing (IgG, IgA, IgM) comprised over 90% of pertussis tests at index encounters, with PCR representing only 5–8%, a pattern present at prior encounters as well. Children aged 5–17 showed a notable shift between prior and index encounters, with PCR increasing from 22.7% to 47.9% and IgG falling from 38.9% to 25.4% (eTable 14).

## Discussion

This study examined patterns of pertussis diagnostic delay across two health systems in South Carolina over a ten-year period spanning the pre- and post-COVID-19 eras, finding significant age-related differences in both the rate and duration of diagnostic delay. Same-day diagnosis was most common among children aged 1–4 years (75.4%), somewhat higher than infants under 1 year (63.6%), and least common among adults 50 and older, nearly half of whom had at least one prior healthcare encounter before pertussis was identified. These findings extend and refine prior work by Evans et al. [18] in several ways: by distinguishing infants from toddlers rather than combining them into a single under-two-years category, by characterizing diagnostic testing patterns not examined in that analysis, and by examining how these patterns were affected by the COVID-19 pandemic, a period not captured in Evans et al.’s claims data.

Differing clinical presentation across the age spectrum is well documented and likely a major contributor to diagnostic delay. While young children typically present with classic pertussis features (paroxysmal cough, inspiratory whoop, post-tussive emesis) that are readily recognized, older adults more often present with prolonged subacute cough easily misattributed to bronchitis or other respiratory illness, contributing to diagnostic delay and underdiagnosis in adults [1, 2, 3, 18, 19, 20]. Our prior encounter data reflect this pattern (eTable 5): acute upper respiratory infection dominated pre-diagnosis encounters in young children, while cough and shortness of breath predominated among adults, underscoring how nonspecific symptoms can obscure recognition until later in the clinical course.

The age-stratified testing patterns add a further dimension to this clinical picture, an aspect not examined by Evans et al. [18], whose risk-factor analysis focused on encounter type and prior antibiotic use rather than diagnostic test selection. PCR predominance in young children, even before diagnosis, reflects providers appropriately pursuing molecular testing in patients with classic early presentations. In contrast, the near-exclusive reliance on serology in adults, present even at pre-diagnosis encounters, suggests that pertussis is not consistently in the differential when providers first evaluate these patients. This distinction carries direct clinical consequences: PCR sensitivity declines substantially after the first two weeks of illness, while serologic IgG responses require two to three weeks to become reliably detectable, creating a diagnostic dead zone in which neither test performs optimally, potentially contributing to the longer delays observed in older adults. By comparison, the shift in children aged 5–17, where PCR use nearly doubled between prior and index encounters, suggests that a deliberate change in testing approach was what ultimately led to diagnosis in this group rather than incidental detection through routine panels.

The adult burden of pertussis in our cohort challenges the persistent perception of pertussis as primarily a pediatric disease. Adults accounted for 48.4% of total cases across both health systems. Delayed diagnosis rates among adults aged 50 and older remained persistently elevated across all three epidemiological periods, suggesting diagnostic challenges predate the pandemic and have not improved during the current resurgence. This pattern is consistent with the global epidemiological shift toward older age groups documented in the post-pandemic resurgence literature [8, 9, 10, 11, 12], and may reflect waning vaccine-derived immunity, as protection from acellular pertussis vaccines has been shown to decline substantially within three to five years of the last dose and may provide little protection beyond seven years [21, 22, 23], compounded by widely acknowledged under-recognition by healthcare professionals of pertussis in older cohorts [20]. The convergence of a susceptible pediatric population and an underrecognized adult reservoir creates a concerning transmission dynamic: delayed adult diagnosis extends the window of unrecognized infectiousness during which household and community spread can occur [7].

Several limitations warrant consideration. First, our lookback approach relies on the presence of respiratory-coded encounters in the EHR prior to the index pertussis date, and some of these encounters may reflect coincidental healthcare utilization rather than true missed diagnostic opportunities. While we applied a 60-day lookback window to minimize this concern, residual misclassification remains possible, particularly in older adults and patients with comorbid respiratory conditions who have higher baseline healthcare utilization. Serologic IgG testing may also yield false positive results in recently vaccinated individuals potentially contributing to misclassification of diagnosis timing in a small number of cases. Finally, neither health system operates as a closed, integrated system; care received elsewhere may not be fully captured in the EHR, meaning prior respiratory encounters outside these systems would not be identified, potentially leading to underestimation of diagnostic delay.

These findings have direct actionable implications. Broadening the differential diagnosis for persistent cough in adults to include pertussis, particularly during periods of elevated community circulation, represents a low-cost intervention with meaningful potential to reduce transmission [19]. Clinicians should also weigh the timing-dependent sensitivity of available tests: prompt molecular testing at first presentation, rather than defaulting to serologic panels, could meaningfully reduce diagnostic delay in adults. The structured EHR-based prior-encounter data documented here may also serve as a substrate for future clinical decision-support tools that flag at-risk patients before confirmation, building on proof-of-concept work showing that machine learning models can support pertussis diagnosis and surveillance [24,25]. Ultimately, the progressive, cross-site age gradient in diagnostic delay identifies adults 50 and older as a high-priority target for clinical awareness initiatives as pertussis reestablishes endemic circulation in the current resurgence.

## Data Availability

Data are not publicly available due to PHI and IRB restrictions.

## Data sharing statement

Data are not publicly available due to PHI and IRB restrictions.

## Author Contributions

Emily Serman (Conceptualization, Formal Analysis, Methodology, Investigation, Writing – original draft, Writing – review and editing), Iromi Jayawardena (Data curation, Formal Analysis, Investigation, Writing – review and editing), Carolina Liskey (Data Curation), Robin LaCroix (Methodology, Writing – review and editing), Alain Litwin (Methodology, Writing – review and editing), Jihad Obeid (Writing – review and editing), Scott Curry (Writing – review and editing), Lior Rennert (Methodology, Supervision, Funding acquisition, Writing – review and editing)

## AI Statement

Claude Sonnet 5 (Anthropic) was used to assist with editing manuscript text and troubleshooting statistical code. The authors reviewed, verified, and take full responsibility for the integrity, accuracy, and content of all text in the final manuscript.

