## Supplemental Materials for "Age-Stratified Patterns of Diagnostic Delay in Pertussis"

Table of Contents

eTable 1. ICD-10 Codes used to identify Pertussis encounters	1

eTable 2. ICD-10-CM Diagnosis Codes for Respiratory Infection-Related Encounters	2-3

eTable 3. Delayed diagnosis rate by age group and COVID-19 period, 60-day lookback window	4

eTable 4. Diagnostic timing by demographic characteristics, 60-day lookback window	4

eTable 5. Diagnoses at prior encounters (encounters before index pertussis date) by age group, 60-day lookback window	5

eTable 6. Co-diagnoses at index encounters (coded on the same data as pertussis diagnosis) by age group	6

eTable 7. Adjusted Hazard Ratios for Same-Day Pertussis Diagnosis From a Discrete-Time Complementary Log-Log Hazard Model	7

eTable 8. Pathogens tested per encounter by care setting and health system, 60-day lookback window	8

eTable 9. Pathogens tested per encounter by age group, 60-day lookback window	8

eTable 10. Pathogens tested per encounter by age group and setting, 60-day lookback window	9

eTable 11. Pathogens tested by diagnostic timing, patient-level, 60-day lookback window	10

eTable 12. Diagnostic timing by index encounter setting	10

eTable 13. Same-day diagnosis rate by age group and index encounter setting	11

eTable 14. Pertussis-specific test modality by age group and encounter type, 60-day lookback window	11

**eTable 1.** ICD-10 codes used to identify Pertussis encounters

| *ICD-10 Code* | *Description* |
| --- | --- |
| A37.00 | Whooping cough due to Bordetella pertussis without pneumonia |
| A37.01 | Whooping cough due to Bordetella pertussis with pneumonia |
| A37.80 | Whooping cough due to other Bordetella species without pneumonia |
| A37.81 | Whooping cough due to other Bordetella species with pneumonia |
| A37.90 | Whooping cough, unspecified species without pneumonia |
| A37.91 | Whooping cough, unspecified species with pneumonia |

**eTable 2.** ICD-10-CM Diagnosis Codes for Respiratory Infection-Related Encounters

| **Condition** | **ICD-10 Codes** |
| --- | --- |
| Parainfluenza | J12.2, J20.4 |
| Adenovirus | A08.2, A85.1, A87.1, B30.0, B30.1, B34.0, B97.0, J12.0, J21.8 |
| Chlamydial pneumonia | J16.0, P23.1 |
| Mycoplasma Pneumoniae | B96.0, J15.7, J20.0 |
| Human Rhinovirus | J20.6 |
| Bordetella parapertussis | A37.10, A37.11 |
| Acute Respiratory Infections (ARI) | A20.2, A22.1, A36.0, A36.1, A36.2, A36.9, A37.10, A37.11, A37.80, A37.81, A37.90, A37.91, A38.0, A38.1, A38.8, A38.9, A48.1, B25.0, B33.4, J00, J01.00, J01.01, J01.10, J01.11, J01.20, J01.21, J01.30, J01.31, J01.40, J01.41, J01.80, J01.81, J01.90, J01.91, J02.0, J02.8, J02.9, J03.00, J03.01, J03.80, J03.81, J03.90, J03.91, J04.0, J04.10, J04.11, J04.2, J04.30, J04.31, J05.0, J05.10, J05.11, J06.0, J06.9, J09.X1, J09.X2, J09.X3, J09.X9, J10.00, J10.01, J10.08, J10.1, J10.2, J10.81, J10.82, J10.83, J10.89, J11.00, J11.08, J11.1, J11.2, J11.81, J11.82, J11.83, J11.89, J12.0, J12.1, J12.2, J12.3, J12.81, J12.82, J12.89, J12.9, J13, J14, J15.0, J15.1, J15.20, J15.211, J15.212, J15.29, J15.3, J15.4, J15.5, J15.61, J15.69, J15.7, J15.8, J15.9, J16.0, J16.8, J17, J18.0, J18.1, J18.2, J18.8, J18.9, J20.0, J20.1, J20.2, J20.3, J20.4, J20.5, J20.6, J20.7, J20.8, J20.9, J21.0, J21.1, J21.8, J21.9, J22 |
| Other Respiratory Infections | J38.4, J39.0, J39.1, J40, J80, J85.0, J85.1, J85.2, J86.0, J86.9, R04.2, R05.1, R05.2, R05.4, R05.8, R05.9, R06.00, R06.02, R06.03, R06.09, R06.1, R06.2, R06.82, R06.89, R06.9, R07.0, R07.1, R07.81, R09.02, R09.1, R09.3, R09.81, R09.82 |
| Chronic Respiratory Conditions | A15.0, A15.4, A15.5, A15.6, A15.7, A15.8, A15.9, A19.0, A19.1, A19.2, A19.8, A19.9, J31.0, J31.1, J31.2, J32.0, J32.1, J32.2, J32.3, J32.4, J32.8, J32.9, J35.01, J35.02, J35.03, J35.8, J35.9, J37.0, J37.1, J41.0, J41.1, J41.8, J42, J43.0, J43.1, J43.2, J43.8, J43.9, J44.0, J44.1, J44.81, J44.89, J44.9, J45.20, J45.21, J45.22, J45.30, J45.31, J45.32, J45.40, J45.41, J45.42, J45.50, J45.51, J45.52, J45.901, J45.902, J45.909, J45.990, J45.991, J45.998, J47.0, J47.1, J47.9, J60, J61, J62.0, J62.8, J63.0, J63.1, J63.2, J63.3, J63.4, J63.5, J63.6, J64, J65, J66.0, J66.1, J66.2, J66.8, J67.0, J67.1, J67.2, J67.3, J67.4, J67.5, J67.6, J67.7, J67.8, J67.9, J68.4, J70.1, J70.3, J81.1, J82.81, J82.83, J84.01, J84.02, J84.03, J84.09, J84.10, J84.111, J84.112, J84.113, J84.115, J84.116, J84.117, J84.170, J84.178, J84.2, J84.81, J84.82, J84.83, J84.841, J84.848, J84.89, J84.9, J93.81, J94.1, J95.3, J96.10, J96.11, J96.12, J96.20, J96.21, J96.22, R05.3 |

**eTable 3.** Delayed diagnosis rate by age group and COVID-19 period, 60-day lookback window

| **Age Group**  (years) | **Pre-COVID-19 (2016-2019)**  n (delayed)/N (total) (%) | **COVID-19 Period (2020-2023)**  n (delayed)/N (total) (%) | **Post-COVID-19 (2024-2026)** n (delayed)/N (total) (%) |
| --- | --- | --- | --- |
| <1 (infant) | 19/48 (39.6) | 4/4 (100) | 13/47 (27.7) |
| 1-4 | 8/43 (18.6) | 7/21 (33.3) | 16/62 (25.8) |
| 5-17 | 23/65 (35.4) | 6/19 (31.6) | 35/97 (36.1) |
| 18-49 | 18/51 (35.3) | 14/37 (37.8) | 36/82 (43.9) |
| 50-64 | 10/20 (50) | 12/23 (52.2) | 26/52 (50) |
| 65+ | 8/19 (42.1) | 14/27 (51.9) | 36/70 (51.4) |

**eTable 4.** Diagnostic timing by demographic characteristics, 60-day lookback window

| **Characteristic** | **N** | **Delayed n/N (%)** | **Median Days to Dx^a^** |
| --- | --- | --- | --- |
| Sex |  |  |  |
| Female | 485 | 202/485 (41.6) | 19 |
| Male | 302 | 103/302 (34.1) | 17 |
| Race |  |  |  |
| Black | 110 | 46/110 (41.8) | 15.5 |
| Other | 113 | 42/113 (37.2) | 13 |
| White | 564 | 217/564 (38.5) | 20 |
| Ethnicity |  |  |  |
| Hispanic | 72 | 31/72 (43.1) | 13 |
| Non-Hispanic | 696 | 270/696 (38.8) | 19 |
| Unknown/Refused | 19 | 4/19 (21.1) | 5 |
| Insurance Type^b^ |  |  |  |
| Commercial/Private | 271 | 118/271 (43.5) | 19 |
| Medicaid | 163 | 60/163 (36.8) | 8.5 |
| Medicare | 113 | 54/113 (47.8) | 27.5 |
| Missing | 31 | 0/31 (0) | N/A |
| Other | 4 | 2/4 (50) | 16 |
| Self-pay/Uninsured | 205 | 71/205 (34.6) | 15 |

a. Median days to diagnosis among delayed patients only
b. Percentages for insurance type calculated among non-missing values

**eTable 5.** Diagnoses at prior encounters (encounters before index pertussis date) by age group, 60-day lookback window

| **Diagnosis** | **N Patients^a^** |
| --- | --- |
| <1 Infant |  |
| Acute upper respiratory infection, unspecified | 20 |
| Cough, unspecified | 12 |
| Acute cough | 6 |
| Vomiting, unspecified | 6 |
| Fever, unspecified | 5 |
| 1-4 |  |
| Acute upper respiratory infection, unspecified | 16 |
| Cough, unspecified | 9 |
| Fever, unspecified | 6 |
| Acute obstructive laryngitis (croup) | 4 |
| Nasal congestion | 4 |
| 5-17 |  |
| Cough, unspecified | 18 |
| Acute upper respiratory infection, unspecified | 14 |
| Fever, unspecified | 7 |
| Unspecified asthma, uncomplicated | 7 |
| Acute cough | 5 |
| Acute sinusitis, unspecified | 5 |
| Moderate persistent asthma, uncomplicated | 5 |
| Other specified cough | 5 |
| 18-49 |  |
| Cough, unspecified | 20 |
| Chronic cough | 10 |
| Shortness of breadth | 10 |
| Acute upper respiratory infection, unspecified | 8 |
| Unspecified asthma, uncomplicated | 8 |
| 50-64 |  |
| Cough, unspecified | 11 |
| Shortness of breath | 11 |
| Acute cough | 10 |
| Pneumonia, unspecified organism | 8 |
| Acute upper respiratory infection, unspecified | 7 |
| 65+ |  |
| Shortness of breath | 10 |
| Cough, unspecified | 9 |
| Acute upper respiratory infection, unspecified | 7 |
| Wheezing | 7 |
| Acute cough | 6 |
| Chronic cough | 6 |
| Diarrhea, unspecified | 6 |

a. Number of distinct patients with this diagnosis at a prior encounter

**eTable 6.** Co-diagnoses at index encounters (coded on the same data as pertussis diagnosis) by age group

| **Diagnosis** | **N Patients^a^** |
| --- | --- |
| <1 Infant |  |
| Cough, unspecified | 14 |
| Vomiting, unspecified | 6 |
| Acute cough | 5 |
| Acute upper respiratory infection, unspecified | 4 |
| Nasal congestion | 4 |
| Subacute cough | 4 |
| 1-4 |  |
| Cough, unspecified | 23 |
| Acute cough | 5 |
| Fever, unspecified | 4 |
| Adenovirus infection, unspecified | 3 |
| Chronic cough | 3 |
| Subacute cough | 3 |
| Vomiting, unspecified | 3 |
| 5-17 |  |
| Cough, unspecified | 18 |
| Acute cough | 13 |
| Subacute cough | 11 |
| Fever, unspecified | 8 |
| Chronic cough | 6 |
| 18-49 |  |
| Cough, unspecified | 17 |
| Chronic cough | 11 |
| Acute cough | 7 |
| Other specified cough | 6 |
| Pneumonia, unspecified organism | 6 |
| Subacute cough | 6 |
| 50-64 |  |
| Cough, unspecified | 13 |
| Chronic cough | 9 |
| Subacute cough | 9 |
| Pneumonia, unspecified organism | 5 |
| Acute cough | 4 |
| 65+ |  |
| Subacute cough | 15 |
| Chronic cough | 10 |
| Cough, unspecified | 10 |
| Pneumonia, unspecified organism | 6 |
| Shortness of breath | 6 |

a. Number of distinct patients with this co-diagnosis at the index encounter. Pertussis-specific diagnoses rows excluded.

**eTable 7.** Adjusted Hazard Ratios for Same-Day Pertussis Diagnosis From a Discrete-Time Complementary Log-Log Hazard Model

| **Characteristic** | **HR^a^** | **95% CI** | **p-value** |
| --- | --- | --- | --- |
| Age group |  |  |  |
| <1 (Infant) | 0.85 | 0.64, 1.13 | 0.3 |
| 1-4^b^ | Ref | Ref | Ref |
| 5-17 | 0.82 | 0.64, 1.05 | 0.11 |
| 18-49 | 0.72 | 0.56, 0.94 | 0.014 |
| 50-64 | 0.58 | 0.43, 0.79 | <0.001 |
| 65+ | 0.56 | 0.38, 0.82 | 0.003 |
| Sex |  |  |  |
| Male | Ref | Ref | Ref |
| Female | 0.87 | 0.75, 1.02 | 0.084 |
| Race |  |  |  |
| White | Ref | Ref | Ref |
| Black | 1.01 | 0.80, 1.26 | >0.9 |
| Other | 0.97 | 0.72, 1.29 | 0.8 |
| Ethnicity |  |  |  |
| Non-Hispanic | Ref | Ref | Ref |
| Hispanic | 0.96 | 0.68, 1.35 | 0.8 |
| Unknown/Refused | 1.74 | 0.96, 2.98 | 0.059 |
| Insurance Type |  |  |  |
| Commercial/Private | Ref | Ref | Ref |
| Self-Pay/Uninsured | 0.99 | 0.79, 1.23 | >0.9 |
| Medicaid | 0.95 | 0.75, 1.21 | 0.7 |
| Medicare | 1.00 | 0.71, 1.41 | >0.9 |
| Other | 1.35 | 0.41, 3.30 | 0.6 |
| Health System |  |  |  |
| Health System A | Ref | Ref | Ref |
| Health System B | 0.79 | 0.66, 0.94 | 0.007 |
| COVID-19 Period |  |  |  |
| Pre-COVID-19 (2016-2019) | Ref | Ref | Ref |
| COVID-19 Period (2020-2023) | 1.00 | 0.79, 1.26 | >0.9 |
| Post-COVID-19 Period (2024-2026) | 0.99 | 0.83, 1.18 | >0.9 |

a. HR = Hazard Ratio. Time-band coefficients (baseline hazard by interval) omitted from this table. Model fit: n=756 patients (7654 person-day observations). Patients with missing insurance data (n=31) were excluded from the model; all 31 had same-day diagnoses, so their exclusion is unlikely to bias estimates of delay-associated factors.
b. Age group 1–4 years was used as the reference category, rather than infants (<1 year), because it had the highest same-day diagnosis rate and the most classically recognized pertussis presentation, providing a clinically meaningful benchmark against which delay in other age groups could be compared.

**eTable 8.** Pathogens tested per encounter by care setting and health system, 60-day lookback window

|  | **Overall** | | **Health System A** | | **Health System B** | |
| --- | --- | --- | --- | --- | --- | --- |
| **Setting** | **N encounters** | **Mean (median)^a^** | **N encounters** | **Mean (median)^a^** | **N encounters** | **Mean (median)^a^** |
| Outpatient | 954 | 4 (2) | 471 | 4.1 (2) | 483 | 3.8 (2) |
| Emergency | 196 | 5.6 (4) | 56 | 4.7 (4) | 140 | 6 (5) |
| Inpatient | 101 | 7.2 (9) | 51 | 7.4 (9) | 50 | 6.9 (9) |

a. Mean (median) number of distinct respiratory pathogens tested per encounter

**eTable 9.** Pathogens tested per encounter by age group, 60-day lookback window

|  | **Overall** | | **Health System A** | | **Health System B** | |
| --- | --- | --- | --- | --- | --- | --- |
| **Age group** | **N encounters** | **Mean (median)** | **N encounters** | **Mean (median)** | **N encounters** | **Mean (median)** |
| <1 (Infant) | 141 | 6.2 (7) | 54 | 6.7 (7) | 87 | 5.9 (9) |
| 1-4 | 139 | 5.9 (7) | 49 | 6 (7) | 90 | 5.9 (4) |
| 5-17 | 285 | 5.1 (5) | 128 | 4.8 (7) | 157 | 5.3 (5) |
| 18-49 | 271 | 3.4 (2) | 146 | 3.9 (2) | 125 | 2.7 (2) |
| 50-64 | 188 | 4.1 (3) | 106 | 3.8 (1.5) | 82 | 4.5 (3) |
| 65+ | 227 | 3.5 (2) | 95 | 3.6 (1) | 132 | 3.4 (2) |

**eTable 10.** Pathogens tested per encounter by age group and setting, 60-day lookback window

|  | **Overall** | | **Health System A** | | **Health System B** | |
| --- | --- | --- | --- | --- | --- | --- |
| **Age group** | **N encounters** | **Mean (median)^a^** | **N encounters** | **Mean (median)^a^** | **N encounters** | **Mean (median)^a^** |
| Outpatient |  |  |  |  |  |  |
| <1 (Infant) | 75 | 5 (4) | 25 | 6.6 (7) | 50 | 4.2 (2) |
| 1-4 | 103 | 5.5 (4) | 40 | 6 (7) | 63 | 5.1 (3) |
| 5-17 | 224 | 4.8 (4) | 110 | 4.7 (7) | 114 | 4.8 (4) |
| 18-49 | 205 | 3 (2) | 120 | 3.6 (2) | 85 | 2.3 (2) |
| 50-64 | 153 | 3.7 (3) | 91 | 3.5 (1) | 62 | 4.1 (3) |
| 65+ | 194 | 3.1 (1) | 85 | 3.2 (1) | 109 | 3 (2) |
| Emergency |  |  |  |  |  |  |
| <1 (Infant) | 45 | 7.3 (9) | 18 | 6.1 (7) | 27 | 8.1 (10) |
| 1-4 | 29 | 6.7 (9) | 8 | 5.8 (5.5) | 21 | 7 (10) |
| 5-17 | 43 | 6 (7) | 9 | 4.6 (3) | 34 | 6.4 (9) |
| 18-49 | 49 | 3.6 (3) | 15 | 3.1 (3) | 34 | 3.8 (4) |
| 50-64 | 17 | 4.2 (4) | 4 | 1.8 (1) | 13 | 5 (4) |
| 65+ | 13 | 5.8 (4) | 2 | 6.5 (6.5) | 11 | 5.6 (4) |
| Inpatient |  |  |  |  |  |  |
| <1 (Infant) | 21 | 8.3 (9) | 11 | 7.9 (7) | 10 | 8.8 (10) |
| 1-4 | 7 | 9.7 (10) | 1 | 7 (7) | 6 | 10.2 (10) |
| 5-17 | 18 | 6.4 (7) | 9 | 5.2 (7) | 9 | 7.7 (9) |
| 18-49 | 17 | 6.6 (9) | 11 | 8.6 (9) | 6 | 3 (2) |
| 50-64 | 18 | 7.1 (8.5) | 11 | 7.5 (9) | 7 | 6.4 (5) |
| 65+ | 20 | 6.2 (7.5) | 8 | 7.1 (9) | 12 | 5.5 (3) |

a. Mean (median) number of distinct pathogens tested per encounter

**eTable 11.** Pathogens tested by diagnostic timing, patient-level, 60-day lookback window

| **Diagnostic timing** | **N patients** | **Mean (median) encounters** | **Mean (median) pathogens^b^** |
| --- | --- | --- | --- |
| Overall^a^ |  |  |  |
| Same-day^c^ | 298 | 1.3 (1) | 4.8 (3) |
| Delayed | 291 | 2.9 (3) | 4.5 (3) |
| Health System A |  |  |  |
| Same-day^c^ | 116 | 1.4 (1) | 5 (7) |
| Delayed | 121 | 3.5 (3) | 4.5 (4) |
| Health System B |  |  |  |
| Same-day^c^ | 182 | 1.3 (1) | 4.7 (2) |
| Delayed | 170 | 2.6 (2) | 4.5 (3) |

a. This table includes 589 of 787 cohort patients (74.8%) who had at least one respiratory pathogen ordered within the 60-day lookback window. The remaining 198 patients (25.2%) had no laboratory testing documented in this period, of whom 184 (92.9%) received a same-day diagnosis.
b. Mean (median) number of distinct pathogens tested across all encounters in the 60-day lookback window
c. Among same-day patients with laboratory data, 64 (21.5%) had more than one encounter on their diagnosis date (Health System A: 30/116, 25.9%; Health System B: 34/182, 18.7%). The majority of Health System A multi-encounter same-day patients (27/30, 90.0%) had multiple outpatient visits on the same date, likely reflecting urgent care and primary care visits occurring on the same day, as both as classified as outpatient in this dataset. Health System B multi-encounter same-day patients showed more diverse encounter combinations, with 15 of 34 (44.1%) involving emergency department or inpatient visits alongside outpatient encounters, consistent with care-escalation on the day of diagnosis. These additional encounters account for the mean number of encounters exceeding 1.0 among same-day patients.

**eTable 12.** Diagnostic timing by index encounter setting

| **Setting^a^** | **Delayed^b^ n/N (%)** | **Same-day n/N (%)** |
| --- | --- | --- |
| Outpatient | 191/456 (41.9) | 265/456 (58.1) |
| Emergency Department | 21/135 (15.6) | 114/135 (84.4) |
| Inpatient | 17/67 (25.4) | 50/67 (74.6) |

a. Index encounter defined as the encounter on the date of pertussis diagnosis
b. Χ^2^(2) = 34.75, p<0.001

**eTable 13.** Same-day diagnosis rate by age group and index encounter setting

| **Age Group^a^** | **Outpatient n (same-day)/N (total) (%)** | **Emergency Department n (same-day)/N (total) (%)** | **Inpatient n (same-day)/N (total) (%)** |
| --- | --- | --- | --- |
| <1 (Infant) | 27/45 (60) | 20/24 (83.3) | 13/20 (65) |
| 1-4 | 54/75 (72) | 27/32 (84.4) | 9/9 (100) |
| 5-17 | 71/105 (67.6) | 28/33 (84.8) | 7/10 (70) |
| 18-49 | 48/94 (51.1) | 25/30 (83.3) | 6/6 (100) |
| 50-64 | 26/59 (44.1) | 9/9 (100) | 7/11 (63.6) |
| 65+ | 39/78 (50) | 5/7 (71.4) | 8/11 (72.7) |

a. Χ^2^(10) = 44.84, p<0.001 for association between age group and index encounter setting

**eTable 14.** Pertussis-specific test modality by age group and encounter type, 60-day lookback window

|  | **PCR%** | **Serology%^a^** | **Other%^b^** |
| --- | --- | --- | --- |
| <1 (Infant) |  |  |  |
| Index | 83.8 | 0.0 | 16.2 |
| Prior | 55.2 | 25.8 | 19.0 |
| 1-4 |  |  |  |
| Index | 72.4 | 16.2 | 11.4 |
| Prior | 70.1 | 26.0 | 3.9 |
| 5-17 |  |  |  |
| Index | 47.9 | 39.6 | 12.5 |
| Prior | 22.7 | 62.5 | 14.7 |
| 18-49 |  |  |  |
| Index | 6.0 | 92.5 | 1.4 |
| Prior | 5.3 | 92.9 | 1.8 |
| 50-64 |  |  |  |
| Index | 7.6 | 92.4 | 0.0 |
| Prior | 5.6 | 93.4 | 1.1 |
| 65+ |  |  |  |
| Index | 5.1 | 93.4 | 1.6 |
| Prior | 9.3 | 90.5 | 0.3 |

a. Serology includes IgG, IgA, and IgM
b. Other comprises unspecified Bordetella pertussis test orders where the testing modality was not documented in the lab name field
